# Housing Tenure and Mental Health Across Adulthood: Evidence from the 1970 British Birth Cohort

**DOI:** 10.64898/2026.11.03.26367190

**Authors:** Rebecca Rhead, Stephani L. Hatch, Jay Das-Munshi, Samantha Davis, Alex Dregan, George B. Ploubidis

## Abstract

**Background:** In the UK, declining home ownership and delayed entry into the housing market have reshaped patterns of housing tenure (whether people own or rent their homes) across adulthood. Home ownership may reflect socioeconomic advantage, but may also provide greater housing security, stability and control, with implications for mental health. Using data from the 1970 British Cohort Study (BCS70), this study examines how home ownership trajectories, defined by whether, when and for how long individuals own their homes, are associated with mental health in midlife.

**Methods:** We analysed data from 8,016 BCS70 participants with information on housing tenure histories and mental health outcomes at age 51. Housing tenure was measured repeatedly from ages 26 to 46 and used to define prespecified patterns of home ownership across adulthood (referred to as ownership regimes), including sustained ownership and never owning. Mental health outcomes at age 51 were psychological distress and positive mental wellbeing. G-estimation of a structural nested mean model was used to estimate contrasts between these ownership regimes while accounting for time-varying confounding affected by prior exposure. Models were estimated in lavaan; non-response weights and full information maximum likelihood were used to address selective attrition and missing data.

**Results:** At age 51, 71% of participants owned their home (23% outright and 48% with a mortgage), while 24% were renting and 5% lived in other arrangements. Sustained home ownership across adulthood was associated with lower psychological distress at age 51 compared with never owning (ATE −0.21, 95% CI −0.47 to −0.03) and higher positive mental wellbeing (ATE 0.56 SD, 95% CI 0.38 to 0.67). In ownership-history regime contrasts, late ownership (first ownership in one’s 40s) was associated with lower psychological distress than never owning (ATE −0.15, 95% CI −0.30 to −0.04), while the estimate for early ownership (first ownership in one’s 30s) was in the same direction but imprecise (ATE −0.12, 95% CI −0.37 to 0.04). For wellbeing, both late and early ownership were associated with higher scores than never owning (ATE 0.17 SD, 95% CI 0.08 to 0.27 and ATE 0.40 SD, 95% CI 0.24 to 0.51, respectively), with the largest association observed for sustained ownership.

**Conclusion:** Home ownership across adulthood (age 26 to 46) was associated with better mental health in midlife (age 51) after accounting for time-varying socio-economic and health-related confounding. Findings suggest that both the timing and persistence of ownership may matter, particularly for positive mental wellbeing, with more sustained exposure to ownership associated with more favourable outcomes.

## Introduction

Housing is widely recognised as a fundamental social determinant of health (1). Aspects of housing such as affordability, quality and residential stability are important for mental health and may differ according to housing tenure. Housing tenure refers to the legal arrangement under which someone occupies a property (e.g. owning or renting a home) and is increasingly recognised as an upstream determinant of mental health (2–5). The relationship between housing tenure and mental health is complex and closely linked to income and wealth. Housing tenure is an established indicator of socioeconomic position (SEP), alongside occupational social class, income, wealth and education (6). However, beyond its role as an indicator of SEP, housing tenure may have distinct implications for mental health through differences in perceived control (7), stability (8), housing quality (9) or stigma associated with particular tenure types (5). These dynamics may vary across generations and life stages, reflecting broader changes in housing policy, labour markets, and social inequality. In the UK, for example, owner-occupation expanded substantially over the late 20th century and was relatively common, whereas access to home ownership has become increasingly constrained for more recent generations. As a result, the meaning and implications of housing tenure, including the relative advantages of ownership compared with renting, may differ across cohorts and over time. This makes it important to examine not only whether housing tenure is associated with mental health, but how changes in tenure across adulthood may shape later mental health outcomes, particularly in the context of increasingly unequal access to secure and affordable housing.

Much of the existing literature investigating housing tenure and mental health is cross-sectional, documenting associations at one point in time without clarifying direction or causality (10). In the UK, cross-sectional studies often find that social renters are more likely to report anxiety and depression than owner-occupiers (11,12). However, longitudinal analyses that yield estimates with a causal interpretation remain relatively scarce. Studies that do explicitly examine how changes in housing tenure affect mental health are “rare but important” (4), with existing research often yielding mixed results.

For example, a recent study using the UK Household Longitudinal Study (13) tracked psychological distress, measured using the General Health Questionnaire, before and after people moved home between 2001 and 2017 using first difference models. They found that moves from renting to home ownership were associated with improvements in mental health. These effects of changing tenure persisted even after adjusting for changes in housing affordability, income, and other factors that vary over time. However, this study only captures short-term changes around the move, and the methods used cannot fully account for time-varying confounders that are affected by earlier tenure. In contrast, analysis of 1991–2009 British Household Panel Survey data compared those who bought their property under Britain’s Right to Buy policy, which enabled social renters to become homeowners, with otherwise similar tenants who stayed renting (4). Using a difference-in-differences design, the authors tracked psychological distress, also measured using the General Health Questionnaire, from two years before to one year after the tenure change and adjusted for shifts in marital status, employment, finances and education. The study found that those who bought their homes experienced no significant reduction in psychological distress compared to those who continued renting, concluding that obtaining ownership delivers no short-term mental health benefit beyond buyers’ pre-existing socio-economic advantage.

International evidence is similarly inconsistent. A 2012 Australian study using longitudinal data from the HILDA (Household, Income and Labour Dynamics in Australia) survey (3) found that while mental health scores differ significantly across housing tenure, these differences were largely attributable to population characteristics such as income, education, employment status, and age. Once these compositional factors were accounted for, there was little evidence of a causal relationship between housing tenure itself and mental health. By contrast, a recent Finnish birth-cohort study (14) tracking everyone born between 1972 and 1975 and linking housing-register, prescription, and hospital records found that, after controlling for earlier mental health treatment, education, income, and occupation, homeowners were significantly less likely than comparable tenants to require psychotropic medication or specialist care. The benefit remained for people with no prior mental health history and was most pronounced among adults who had grown up in owner-occupied housing or lone-parent families, suggesting a protective effect of homeownership. Length of time in ownership added little, indicating that tenure status itself matters most.

Against a backdrop of declining housing affordability and widening inequalities in access to home ownership in the UK (15), there is a gap in our understanding of the impact of housing tenure on mental health outcomes across the life course. Such knowledge is essential for motivating housing policy changes to address widening health inequalities in the UK. We used data from the 1970 British Cohort Study (BCS70), a birth cohort with prospectively collected information spanning more than five decades. Since housing tenure, mental health, and their shared determinants are measured repeatedly from early life onwards, BCS70 allows tenure histories to be observed across the life course and their effects on mental health to be examined over time. Focusing on this cohort is particularly useful because cohort members formed households during the late 1980s and 1990s, a period marked by recessions, rising inequality and shifts in housing policy that may have shaped opportunities for home ownership and had lasting implications for mental health in midlife (16).

Specifically, this study aims to provide answers to the following research questions:

1. How do different housing tenure trajectories across adulthood relate to mental health outcomes in midlife?
2. What is the association of sustained exposure to home ownership across adulthood, compared with never owning, on mental health in midlife?

## Methods

### Data

We used data from the 1970 British Cohort Study (BCS70), a national birth cohort study that follows the lives of individuals born in England, Scotland, and Wales in a single week in 1970 (17). This involved 17,196 babies at baseline, with 11 follow-up sweeps conducted when the cohort members were aged 5, 10, 16, 26, 30, 34, 38, 42, 46 and 51.

At age 51, 12,368 participants were invited to participate (71.9% of original), and 8,016 (64.8%) were interviewed (18). Trained interviewers made initial contact and conducted core interviews, including a cognitive assessment module. Among other factors, BCS70 collects information on participants’ health, educational, social, and physical development. Although attrition in BCS70 has been socio-demographically patterned, particularly higher among men and participants from lower socio-economic backgrounds (19), extensive work shows that through application of the survey’s inverse-probability non-response weights and full information maximum likelihood estimation, the cohort’s baseline representativeness can be fully restored (20–23).

### Measures

#### Housing tenure and home ownership

Housing tenure was assessed at ages 26, 30, 34, 42, 46, and 51 (the age 38 sweep was excluded because it collected a more limited range of measures). Participants reported their housing situation, which was grouped into four categories: owned outright, owned with a mortgage, rented (including both social housing and renting from private landlords), and other arrangements. The ‘other’ category included living rent-free, squatting, and other non-standard arrangements.

Housing tenure was also converted into a binary exposure indicating home ownership, with participants classified as owners if they owned their home outright or with a mortgage, and as non-owners if they were renting or in other arrangements. Four prespecified longitudinal groups (referred to as ownership-history regimes) were defined to represent patterns of exposure to home ownership across adulthood: never owning, defined as no home ownership at any observed adult exposure sweep; late ownership, defined as first entering home ownership at age 42 or 46 and remaining an owner at subsequent exposure sweeps; early ownership, defined as first entering home ownership at age 30 or 34 and remaining an owner at subsequent exposure sweeps; and always owning, defined as home ownership at every observed adult exposure sweep.

#### Mental health outcomes

Psychological distress was assessed at each sweep using the Malaise Inventory. Cohort members completed the 9 yes/no items (e.g., “Do you often feel miserable and depressed?”). Affirmative responses are summed to give a total score from 0 to 9, with higher scores reflecting greater distress. The Malaise Inventory has shown good internal consistency in UK population studies (Cronbach’s α ≈ 0.80) and convergent validity against clinically diagnosed depression and anxiety (24,25). It has also demonstrated measurement invariance over time, with Item Response Theory analyses indicating greater measurement precision at moderate to high levels of psychological distress (26).

Positive mental wellbeing was assessed using the 14-item Warwick–Edinburgh Mental Well-Being Scale (WEMWBS) at ages 42 and 46, and the 7-item Short Warwick–Edinburgh Mental Well-Being Scale (SWEMWBS) at age 51. Because the wellbeing measure differed across sweeps, wellbeing scores were standardised before analysis. The age 51 standardised SWEMWBS score was used as the wellbeing outcome, while standardised wellbeing scores at ages 42 and 46 were included as prior wellbeing covariates. Because wellbeing scores were standardised, estimated effects on wellbeing are expressed in standard deviation units rather than raw scale points. The WEMWBS and SWEMWBS have shown good psychometric properties in UK population samples and are recommended for population-based surveys and cohort studies (27).

#### Covariates

Covariates were selected based on the hypothesised causal relationships shown in the directed acyclic graph (DAG; **Figure 1**), to capture pre-exposure and time-varying factors that may confound the relationship between housing tenure and mental health. Time-varying confounders were measured across the adult sweeps where available and included:

**Figure 1:**
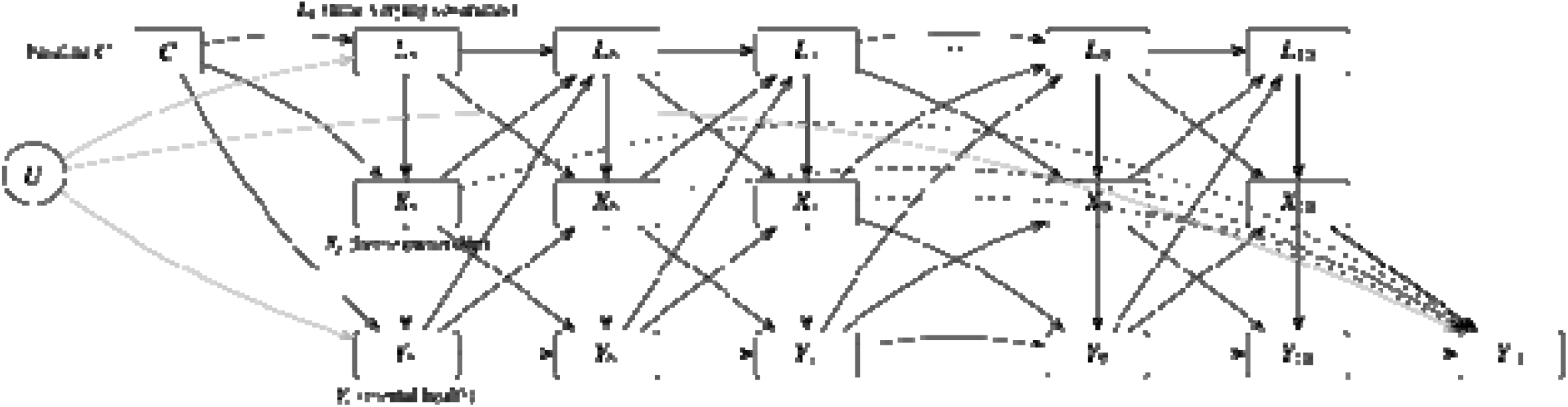
Directed acyclic graph (DAG) illustrating the g-estimation framework used to examine the causal effect of housing tenure across multiple BCS70 sweeps. Socio-economic and health-related factors are accounted for as time-varying covariates. A circular node denotes unmeasured factor(s) U. For visual clarity, arrows emanating from U are shown in grey. Dashed arrows indicate the treatment (blip) effects of housing tenure at each sweep on the final mental health outcome, as specified in the structural nested mean model.

- Self-rated general health - coded as excellent/very good, good, fair, and poor/very poor.
- Occupational social class - harmonised across sweeps using a five-category occupational class measure: professional/managerial, skilled non-manual, skilled manual, partly skilled, and unskilled.
- Weekly income or earnings - converted to a weekly measure where required and log-transformed for analysis.
- Prior mental health - included where available. For psychological distress models, prior Malaise Inventory scores were included at earlier adult sweeps. For wellbeing models, prior standardised WEMWBS scores were included at ages 42 and 46.

Time-invariant early-life covariates were selected to capture factors preceding adult housing tenure that may influence both subsequent housing trajectories and mental health, informed by the hypothesised causal relationships shown in the DAG (Figure 1) and previous analyses of BCS70 data (28–30). These included:

- Parental social class at birth - measured using Registrar General Social Classes I–V.
- Parental housing tenure at age 10 - measured as the cohort member’s family housing tenure in childhood.
- Cognitive ability at age 10 - derived from a principal component analysis of age 10 cognitive test scores, including reading and mathematics measures.
- Internalising symptoms at age 16 - measured using the total Rutter A scale.

### Analyses

This study addressed two related research questions. The first examined the association between sustained home ownership across adulthood and mental health in midlife, accounting for time-varying socio-economic and health-related confounding, as well as early-life characteristics. The second examined how different ownership-history regimes across adulthood were associated with mental health outcomes at age 51. G-estimation was used to address both research questions within a causal inference framework, while accounting for time invariant and time-varying confounding. This approach is well suited to longitudinal settings in which both exposures and confounders vary over time, and where confounders may themselves be influenced by prior exposure (31). G-estimation was implemented using a structural nested mean model (SNMM), which estimates contrasts in potential outcomes under alternative exposure histories. Interpretation of these estimates relies on the usual assumptions of consistency, positivity, and no unmeasured confounding conditional on the measured covariate history.

To address the first research question, exposure was defined as home ownership at observed adult sweeps from age 26 to age 46. The estimand (the causal effect of interest) represented the average difference in mental health outcomes at age 51 under a static regime in which all individuals were assumed to own their home at every observed adult exposure sweep, compared with a regime in which none were assumed to own their home at any observed adult exposure sweep, conditional on measured confounders. To address the second research question, prespecified ownership-history regimes were constructed and compared within the same SNMM framework. These represented never owning, late ownership, early ownership, and always owning, as defined above. Each ownership-history regime was compared with never owning to assess whether mental health outcomes at age 51 differed according to the timing and persistence of home ownership across adulthood.

These SNMMs were estimated using the lavaan package (32), following procedures outlined by Loh and Ren (33). Model specification was informed by the DAG displayed in **Figure 1**. Adjustment included time-varying occupational social class, weekly income or earnings, self-rated general health, and prior mental health where available, as well as early-life characteristics including parental social class at birth, parental housing tenure at age 10, cognitive ability at age 10, and internalising symptoms at age 16.

All models were estimated in lavaan using robust maximum likelihood (MLR), which provides robust standard errors and scaled test statistics to account for departures from normality, including skewness in the Malaise Inventory scores. The analytic sample was restricted to sweep 11 participants with non-missing age-51 outcome data, a valid sweep 11 non-response weight, and at least one observed adult home ownership measure. Sweep 11 non-response weights supplied with the BCS70 data were applied to account for differential participation at age 51. These inverse-probability weights were derived from models predicting response using demographic, socio-economic, health, and behavioural characteristics measured at earlier sweeps (18). Confidence intervals for ownership-regime contrasts were obtained using a nonparametric percentile bootstrap. All analysis was conducted in R version 4.2.2 (34).

## Results

At age 51, sociodemographic and health characteristics of BCS70 participants varied by housing tenure (as shown in Table 1). Among those with a mortgage, 57% were in higher or intermediate occupational classes, compared with 29% of renters. Median household income was also highest among participants with a mortgage (£965 per week) and lowest among renters (£416) and those in the ‘other’ category (£406). Economic activity reflected this, with 92% of participants with a mortgage being in employment and 84% of outright owners, compared with 62% of renters and 67% of those in the ‘other’ category.

**Table 1:** Sociodemographic and health characteristics of BCS70 participants by tenure at age 51.

| Characteristic | Overall = 8016 | **Own outright**<br>N = 2102 (23%)<sup>1</sup> | Own with mortgage<br>N = 4238 (48%)<sup>1</sup> | **Rent**<br>N = 1348 (24%)<sup>1</sup> | **Other** <sup>a</sup><br>N = 328 (4.8%)<sup>1</sup> |
|---|---|---|---|---|---|
| **Birth sex** |  |  |  |  |  |
| Male | 3,805 (49.4%) | 911 (44.9%) | 2,103 (50.2%) | 628 (50.1%) | 163 (58.4%) |
| Female | 4,211 (50.6%) | 1,191 (55.1%) | 2,135 (49.8%) | 720 (49.9%) | 165 (41.6%) |
| **Psychological distress** <sup>b</sup> |  |  |  |  |  |
| Low psychological distress (0-3) | 6,173 (79.8%) | 1,709 (84.4%) | 3,390 (84.9%) | 836 (65.4%) | 238 (76.2%) |
| High psychological distress (4+) | 1,253 (20.2%) | 283 (15.6%) | 544 (15.1%) | 366 (34.6%) | 60 (23.8%) |
| **Wellbeing score** <sup>c</sup> | 26.0 (22.0, 28.0) | 26.0 (23.0, 28.0) | 26.0 (23.0, 28.0) | 24.0 (20.0, 28.0) | 25.0 (22.0, 28.0) |
| General health (self-reported) |  |  |  |  |  |
| Excellent/Very good | 3,791 (41.7%) | 1,104 (50.3%) | 2,200 (48.2%) | 358 (23.0%) | 129 (29.6%) |
| Good | 2,529 (31.0%) | 622 (29.0%) | 1,390 (33.3%) | 423 (28.2%) | 94 (30.4%) |
| Fair | 1,171 (17.9%) | 273 (14.9%) | 506 (14.0%) | 325 (27.0%) | 67 (24.2%) |
| Poor/Very poor | 468 (9.4%) | 90 (5.7%) | 121 (4.4%) | 225 (21.7%) | 32 (15.9%) |
| **NS-SEC** <sup>d</sup> |  |  |  |  |  |
| Higher managerial, administrative and professional occupations | 3,536 (49.1%) | 891 (49.3%) | 2,252 (56.6%) | 304 (29.3%) | 89 (34.8%) |
| Intermediate occupations | 801 (11.4%) | 204 (11.0%) | 468 (11.8%) | 98 (10.1%) | 31 (14.2%) |
| Small employers and own account workers | 940 (14.8%) | 276 (17.1%) | 486 (14.1%) | 136 (13.3%) | 42 (19.0%) |
| Lower supervisory and technical occupations | 410 (7.1%) | 113 (7.4%) | 193 (5.6%) | 84 (10.9%) | 20 (8.0%) |
| Semi-routine and routine occupations | 981 (17.6%) | 235 (15.1%) | 407 (12.0%) | 286 (36.4%) | 53 (24.0%) |
| Average weekly household income | 769 (449, 1,162) | 812 (511, 1,175) | 965 (679, 1,378) | 416 (284, 683) | 406 (276, 825) |
| **Economic activity** |  |  |  |  |  |
| Employed | 6,968 (81.5%) | 1,798 (83.9%) | 3,967 (91.8%) | 955 (61.7%) | 248 (66.8%) |
| Unemployed | 109 (2.0%) | 23 (1.2%) | 27 (0.7%) | 48 (4.8%) | 11 (5.9%) |
| Sick/disabled | 367 (8.7%) | 62 (4.6%) | 57 (2.8%) | 208 (21.9%) | 40 (19.5%) |
| Retired/Home/Student/Other | 565 (7.8%) | 216 (10.3%) | 186 (4.7%) | 135 (11.6%) | 28 (7.8%) |

<sup>1</sup>n (unweighted) (%); Median (IQR)
‘Other’ housing tenure = living rent-free (e.g., with relatives or friends), squatting, or other arrangements
Psychological distress measured using Malaise Inventory
Wellbeing measured using WEMWBS at ages 42 and 46 and SWEMWBS at age 51
NS-SEC = National Statistics Socio-economic Classification

Being unable to work due to sickness or disability was more common among renters (22%) and those in the ‘other’ group (20%) than among homeowners (5% of outright owners and 3% of mortgage holders). Poor or very poor self-rated health was reported by 4% of participants with a mortgage and 6% of outright owners, compared with 22% of renters and 16% of those in the ‘other’ category. High psychological distress was also more prevalent among non-owners, observed in 35% of renters compared with 15% of mortgage holders and 16% of outright owners.

Figure 2 illustrates patterns of housing tenure across adulthood, displaying transitions from ages 26 to 51 among those who took part in the age 51 sweep. At age 26, renting was the most common tenure (48%), with 46% of participants owning their home outright or with a mortgage. By age 30, home ownership had increased to 68%, rising to 78% by age 34. Ownership remained the dominant tenure type throughout midlife, with the proportion owning outright increasing from 2% at age 26 to 26% at age 51. However, transitions were not always towards home ownership. Some participants moved from ‘other’ arrangements directly into ownership, while others moved from owning with a mortgage into renting between observed sweeps.

**Figure 2.**
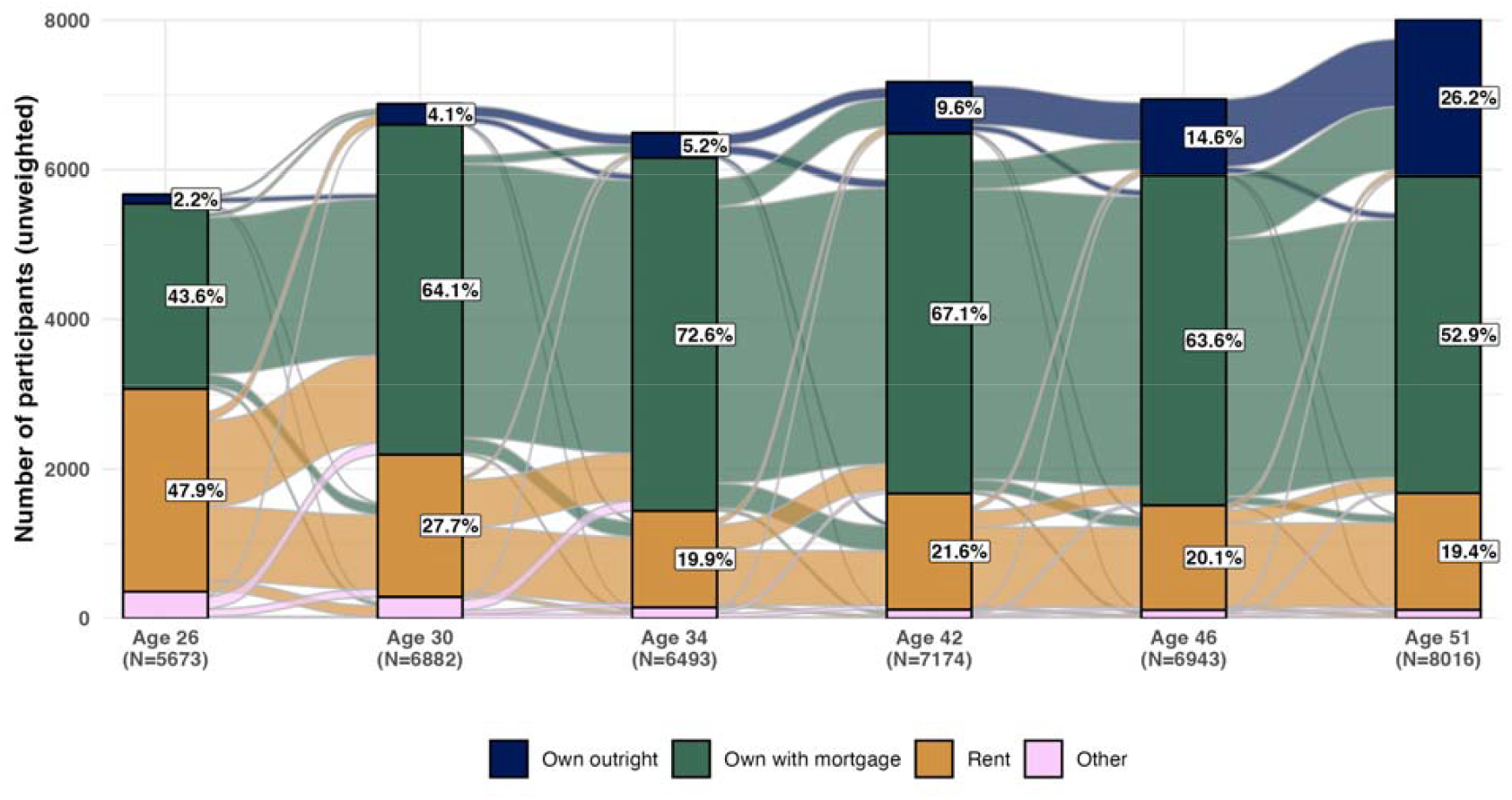
Housing tenure transitions across adulthood among BCS70 participants included in the age 51 analytic sample. The alluvial plot shows movement between housing tenure categories measured at ages 26, 30, 34, 42, 46 and 51. Strata heights represent the number of participants in each tenure category at each age, and flows represent observed transitions between categories across sweeps. Full counts and percentages by housing tenure at each age are provided in Supplementary Table A.

### Sustained home ownership on mental health

Using g-estimation of a structural nested mean model to account for time-varying socio-economic and health-related confounding, sustained home ownership across adulthood was associated with lower psychological distress at age 51 compared with never owning. Under a static regime in which all individuals were assumed to own their home at each observed adult exposure sweep from ages 26 to 46, psychological distress at age 51 was estimated to be 0.21 points lower than under a regime of never owning (ATE −0.21, 95% CI −0.47 to −0.03). Sustained home ownership was also associated with higher wellbeing at age 51, with wellbeing estimated to be 0.56 standard deviations higher under the always-owning regime than under the never-owning regime (ATE 0.56, 95% CI 0.38 to 0.67). Figures 3 and 4 present the ownership-history regime contrasts for psychological distress and wellbeing, respectively, including the sustained-ownership contrast reported here.

**Figure 3.**
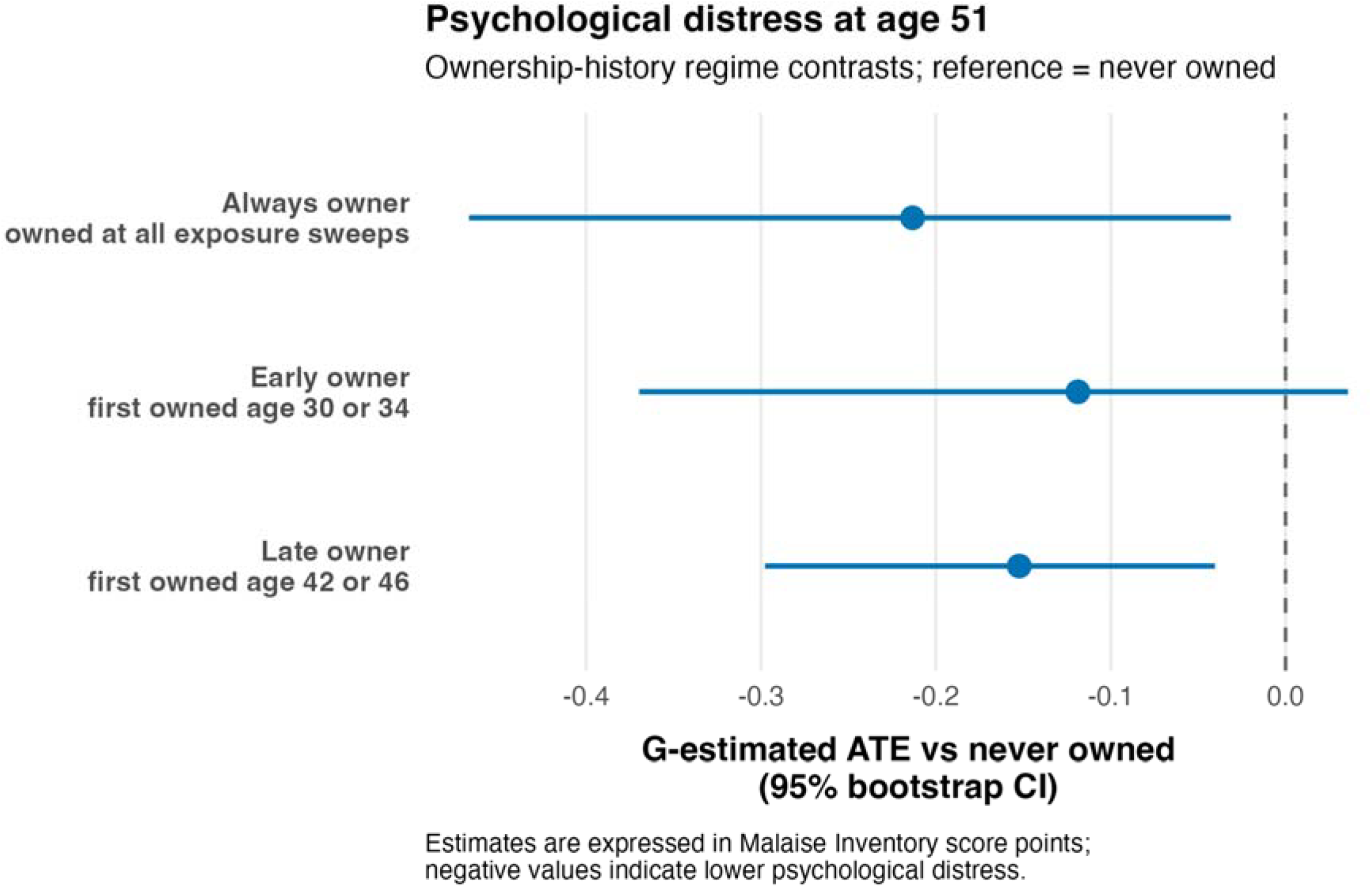
Ownership-history regime contrasts for psychological distress at age 51. Estimated differences in psychological distress at age 51 under prespecified ownership-history regimes compared with never owning. Points show g-estimated average treatment effects (ATEs) from a structural nested mean model; horizontal lines show 95% percentile bootstrap confidence intervals. The reference regime was never owning at the observed adult exposure sweeps. “Late owner” denotes first ownership at age 42 or 46, “early owner” denotes first ownership at age 30 or 34, and “always owner” denotes ownership at all observed adult exposure sweeps. Estimates are expressed in Malaise Inventory score points; negative values indicate lower psychological distress relative to never owning.

**Figure 4.**
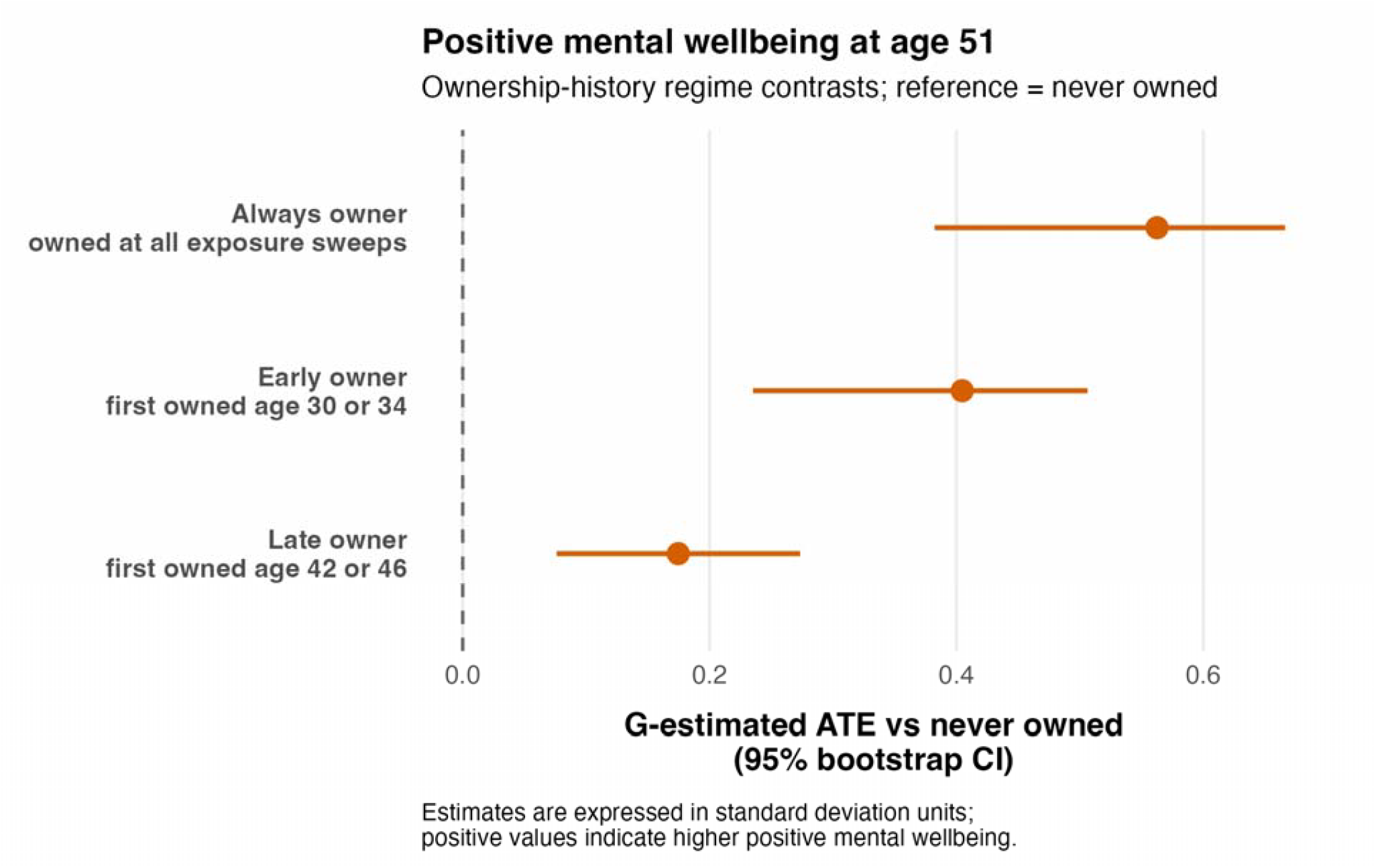
Ownership-history regime contrasts for positive mental wellbeing at age 51. Estimated differences in positive mental wellbeing at age 51 under prespecified ownership-history regimes compared with never owning. Points show g-estimated average treatment effects (ATEs) from a separate structural nested mean model; horizontal lines show 95% percentile bootstrap confidence intervals. The reference regime was never owning at the observed adult exposure sweeps. “Late owner” denotes first ownership at age 42 or 46, “early owner” denotes first ownership at age 30 or 34, and “always owner” denotes ownership at all observed adult exposure sweeps. Estimates are expressed in standard deviation units; positive values indicate higher positive mental wellbeing relative to never owning.

### Tenure Regime Contrasts

To examine whether the timing and persistence of home ownership were associated with mental health at age 51, prespecified ownership-history regimes were compared with never owning within the same g-estimation framework. For psychological distress (Figure 3), late ownership was associated with lower distress at age 51 compared with never owning (ATE −0.15, 95% CI −0.30 to −0.04). Early ownership was also associated with lower distress, although the confidence interval included the null (ATE −0.12, 95% CI −0.37 to 0.04). For wellbeing (Figure 4), all ownership-history regimes were associated with higher scores at age 51 compared with never owning. Late ownership was associated with wellbeing 0.17 standard deviations higher than never owning (ATE 0.17, 95% CI 0.08 to 0.27). Early ownership was associated with wellbeing 0.40 standard deviations higher (ATE 0.40, 95% CI 0.24 to 0.51). This pattern was consistent with a gradient in wellbeing according to earlier and more sustained exposure to home ownership.

Additional pairwise contrasts using early ownership as the reference are presented in Supplementary Table B. Estimated psychological distress was lower under the always-owner regime than under the early-owner regime (mean difference −0.09, 95% CI −0.22 to −0.01), while there was little evidence of a difference between the late-owner and early-owner regimes (−0.03, 95% CI −0.16 to 0.09). For positive mental wellbeing, estimated wellbeing was higher under the always-owner regime than under the early-owner regime (0.16 SD, 95% CI 0.09 to 0.20), but lower under the late-owner regime than under the early-owner regime (−0.23 SD, 95% CI −0.30 to −0.13).

## Discussion

This study provides new evidence on how home-ownership histories across adulthood are associated with mental health in midlife. Using g-estimation to address measured time-varying socioeconomic and health-related confounding, we found that ownership throughout the observed adult sweeps was associated with lower psychological distress and higher positive mental wellbeing at age 51 compared with never owning. Analyses of prespecified ownership histories indicated that the pattern differed by outcome. For wellbeing, pairwise comparisons supported a graded pattern: always owners had higher estimated wellbeing than early owners, who had higher wellbeing than late owners, while never owners had the lowest estimated wellbeing. For psychological distress, the pattern was less consistent: always owners had slightly lower estimated distress than early owners, but early and late owners did not clearly differ. Taken together, these findings suggest that the timing and persistence of home ownership may be particularly relevant to positive mental wellbeing. They also support conceptualising housing tenure as more than a marker of socioeconomic position: it may additionally capture material assets and aspects of housing security, stability and control. However, these potential pathways were not directly tested and remain hypotheses for future research.

The 1970 cohort occupies an important position in generational changes in housing. Improvements in home ownership seen across earlier post-war generations began to stall among those born in the 1970s, with subsequent cohorts experiencing substantially lower rates of ownership at comparable ages (35). Home ownership may therefore capture more than current socio-economic position. Over time, ownership can allow the accumulation of housing wealth and provide an asset that may offer some protection from financial precarity, while also providing greater residential security and control over the home (36). These potential benefits may become increasingly important as people move through midlife and towards retirement. However, these mechanisms were not directly examined in the present study and should be tested in future research.

These findings are relevant to the concept of Generational Health Drift (37). Evidence from British birth cohort comparisons indicates that more recent generations have experienced poorer health at comparable ages, with particularly consistent differences in psychological distress in adulthood (37). The 1970 cohort has repeatedly been found to report higher psychological distress than earlier-born British cohorts. Our study cannot establish whether differences in home ownership explain these between-cohort differences, but it identifies sustained home ownership as one plausible life-course exposure that could contribute to them and could be examined directly in future cross-cohort analyses.

Our findings extend previous longitudinal research, which has largely focused on short-term changes following entry into home ownership and has produced mixed results (3,4,13,14). In contrast, our results suggest that the timing and persistence of ownership across adulthood may be important, particularly for wellbeing. This supports considering housing tenure as a cumulative life-course exposure, potentially reflecting both sustained housing security and the accumulation of housing wealth over time. In later-born cohorts, delayed entry into ownership and more fragmented housing trajectories (16) may therefore reduce exposure to these potential benefits, providing one possible structural pathway relevant to the Generational Health Drift (37).

### Implications

Our findings suggest that the mental health relevance of housing may extend beyond entry into home ownership to the timing, affordability and persistence of ownership across adulthood. Several mechanisms could potentially explain these associations. Sustained ownership may provide greater residential security, stability and control, while enabling the accumulation of housing wealth and reducing exposure to involuntary moves. However, ownership may also create financial strain through mortgage repayments, maintenance costs and changes in interest rates, particularly when housing costs account for a substantial proportion of household income. These mechanisms were not directly examined in this study and should be investigated in future research. Accordingly, our findings should not be interpreted as evidence that promoting home ownership alone will improve mental health. Instead, they emphasise the potential importance of sustained access to stable and affordable housing across tenures, including genuinely affordable housing supply and greater security in the private rented sector.

These findings may also have implications for future generations. If later-born cohorts spend longer in insecure or unaffordable housing, enter home ownership later, and accumulate less housing wealth, this could contribute to the generational differences in mental health described by Generational Health Drift (37). This should be examined directly in future cross-cohort analyses to assess whether differences in housing trajectories account for part of the mental health gap between generations.

Future research should also examine adopt an intersectional approach and investigate whether these associations differ by sex, ethnicity, social class and other markers of socio-economic disadvantage. Access to stable home ownership is itself socially patterned, and understanding who is most affected by changes in housing trajectories will be important for identifying whether housing contributes to inequalities in mental health within as well as between generations.

### Strengths and Limitations

This study draws on a large, long-running birth cohort with repeated measures of housing tenure, socio-economic circumstances and mental health across adulthood, allowing us to examine long-term patterns of exposure rather than relying on cross-sectional associations. The application of g-estimation within a structural nested mean model allowed us to account for time invariant but also time-varying confounders that may themselves be affected by earlier housing tenure. In addition, examining both psychological distress and positive wellbeing provided complementary perspectives on mental health in midlife.

Several limitations should nevertheless be considered. Although we adjusted for a wide range of early-life and time-varying socio-economic and health-related factors, residual confounding from unmeasured characteristics, such as personality or aspects of financial security not captured in the cohort, cannot be ruled out. Causal interpretation of the SNMM estimates therefore depends on assumptions including no unmeasured confounding, positivity, consistency and correct model specification. However, the richness of BCS70 and availability of repeated measures across adulthood enabled adjustment for a broad range of factors likely to influence both housing tenure and mental health.

Housing tenure was dichotomised into owner and non-owner status to operationalise the time-varying exposure within the binary SNMM framework used in this study and to define interpretable ownership-history regimes across adulthood. This provided a consistent measure across repeated sweeps but necessarily obscured potentially important differences between outright and mortgaged ownership, and between private renting, social renting and other arrangements. The analysis also did not directly capture several mechanisms that may distinguish these housing circumstances, including housing wealth, mortgage or rental burden, affordability, security of tenure and control over the home. These remain plausible pathways linking housing tenure to mental health but were not directly tested in the present study.

Measurement of prior mental health also differed between the two outcomes. Psychological distress was available repeatedly across the adult sweeps, whereas prior wellbeing was available only at ages 42 and 46. Adjustment for pre-existing wellbeing was therefore more limited, although including wellbeing at both sweeps provided some control for differences preceding the age-51 outcome. Finally, the analysis is restricted to a single cohort born in 1970, so the findings may not generalise directly to generations experiencing different housing markets or policy environments. However, the repeated prospective measurements in BCS70 provide unusually detailed information on housing and mental health across more than two decades of adult life.

## Conclusion

Sustained exposure to home ownership across adulthood was associated with better midlife mental health in the 1970 British Cohort Study, with particularly clear evidence of a graded association with wellbeing according to the timing and persistence of ownership. These findings suggest that housing tenure may be relevant to mental health across the life course, potentially through greater residential security and control, the accumulation of housing wealth, or a combination of these processes. Future research should seek to disentangle these mechanisms and examine whether changing housing trajectories contribute to generational differences in mental health. In the context of declining and delayed access to home ownership, improving access to stable, secure and affordable housing should be considered an important part of the public mental health agenda.

## Data Availability

Data for BCS70 (SN 8547) are available through the UK Data Service.

https://datacatalogue.ukdataservice.ac.uk/series/series/200001#abstract

## Acknowledgements

BCS70 is supported by the Centre for Longitudinal Studies, Resource Centre 2015-20 grant (ES/M001660/1) and a host of other co-funders.

S.L.H. and R.R. are supported by Wellcome [308556/Z/23/Z], the Economic and Social Research Council Centre for Society and Mental Health at King’s College London [ES/S012567/1]. S.L.H, RR, SD, JD and AD are also supported by UK Research and Innovation [MR/Y030788/1] as part of Population Health Improvement UK (PHI-UK), a national research network that seeks to transform health and reduce inequalities through change at the population level. S.L.H. also receives funding from Impact on Urban Health part of Guy’s & St Thomas’ Foundation [EIC210605 and EIC221208], the Swedish Research Council [2023-05959] and Wellcome [228117/Z/23/Z & 223486/Z/21/Z]. RR acknowledges funding from the Medical Research Council (MR/W021277/1). GP received funding from ESRC [ES/W013142/1]. AD work is funded by NIHR (NIHR163394). The funders had no involvement in study design, data collection, analysis, interpretation or the decision to submit for publication. The views expressed are those of the author(s) and not necessarily those of the funders.

## Supplementary

**Supplementary Table A.**
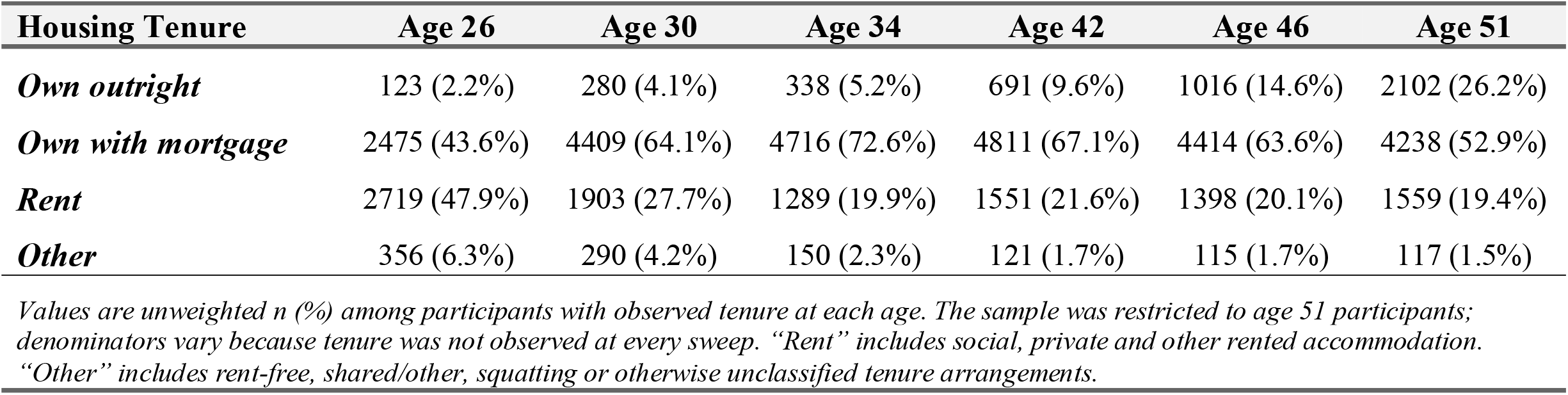
Distribution of housing tenure across adulthood among BCS70 participants included in the age 51 analytic sample.

**Supplementary Table B.** Pairwise contrasts using early ownership as reference.

| Outcome | Comparison | Mean difference | 95% Confidence Intervals |
|---|---|---|---|
| ***Psychological distress*** | Never owner vs early owner | 0.12 | -0.04 to 0.37 |
|  | Late owner vs early owner | -0.03 | -0.16 to 0.09 |
|  | Always owner vs early owner | -0.09 | -0.22 to -0.01 |
| ***Positive mental wellbeing*** | Never owner vs early owner | -0.40 | -0.51 to -0.24 |
|  | Late owner vs early owner | -0.23 | -0.30 to -0.13 |
|  | Always owner vs early owner | 0.16 | 0.09 to 0.20 |

*Estimates represent the mean outcome difference for the named regime minus the early-owner regime. Negative values indicate lower psychological distress; positive values indicate higher positive mental wellbeing. Psychological distress is expressed in Malaise Inventory score points. Positive mental wellbeing is expressed in standard deviation units. Never owner indicates no ownership at observed adult sweeps; late owner indicates first ownership at age 42 or 46; early owner indicates first ownership at age 30 or 34; always owner indicates ownership at all observed adult sweeps.*

